# Standardization of *Trypanosoma cruzi* DNA extraction and purification protocol from samples collected on Whatman 903 filter paper to Chagas disease diagnosis

**DOI:** 10.1101/2020.07.16.20153916

**Authors:** LS Jurado Medina, G Ballering, M Bisio, RM Ojeda, J Altcheh

## Abstract

Chagas disease (CD) caused by the parasite *Trypanosoma cruzi*, belongs to the so-called neglected diseases group. In Argentina about 1,500 children are born with congenital Chagas disease per year. The diagnosis of CD in the newborn relies on the ability to detect parasites in the blood by microscopic observation, as the serological tests are ruled out because of the presence of maternal antibodies. CD treatment is more effective during the acute phase of infection. Early diagnosis and treatment of the disease is thus very important. The Argentinian National Program for early detection of metabolic diseases uses Whatman903 filter paper for blood sampling. This type of sample collection presents many advantages as the use of low blood volumes, minimal biological risk, and easy storage and transportation. The objective of the study was to evaluate the conservation efficiency of blood samples on filter paper in order to access good sensitivity on qPCR results for the detection of *T. cruzi*. To standardize the procedure, negative samples of blood were infected artificially with serial dilutions of trypomastigotes forms of *T. cruzi* from the TcVI strain obtained by cell culture in Vero cells. Concentrations between 50000 and 5 parasites/mL were prepared and loaded in filter paper for analysis. DNA extraction was conducted by the QIAamp DNA Mini Kit from QIAGEN. For qPCR, a method based on TaqMan technology was used, with a multiplex reaction for quantification of *T. cruzi* satellite DNA and an internal amplification control (IAC). The detection limit found from our results was 400 parasites/mL, demonstrating that this method could be a reliable option for the diagnosis of congenital CD by the detection of *T. cruzi* in blood collected in filter paper.

## Introduction

Chagas disease (CD) is caused by the parasite *Trypanosoma cruzi* (*T. cruzi*) affecting more than 6 million people only in Latin America (WHO 2018). Recently, CD has been recognized as a global health concern due to the population mobility that creates opportunities for the spread and the establishment of the disease (Schmunis et al 2010).

*T. cruzi* is transmitted to humans mainly by direct contact with vector feces, by blood transfusion and vertical transmission. Currently the vertical transmission remains the most common infection route (Carlier et al., 2019, Moscatelli et al 2013). The overall transmission rate is about 1-12% with an average of 5% (Bustos et al. 2019). Commonly used methods to diagnose CD in newborns are complicated, the technique recommended by the WHO (Carlier et al., 2019). is direct observation under the microscope (Microhematocrit test) but has limited sensitivity.

In newborns, CD treatment is highly effective and is well tolerated with few adverse events (Altcheh et al. 2001, Moscatelli et al. 2019), and was also shown to stop disease progression (Viotti et al. 2011). Early detection of *T. cruzi* is a key strategy to reduce the disease impact especially in congenital cases (OMS, WHO 2019).

Molecular diagnostic techniques have proven to be an alternative to conventional diagnostic tests. T. *cruzi*- qPCR has shown high reliability and good sensitivity (0.5 parasites/mL) in blood but require toxic preservative agents (Guanidine) (Duffy et al. 2013) and strict transport conditions. Thus, there is an urgent need for the development of a reliable and sensitive input-required screening system for *T. cruzi* infection that allows early diagnosis.

Filter paper is widely used as a standardized blood carrier for governmental public health programs including the Argentinean National Program for Screening of Metabolic Diseases (msal.gob.ar 2011)^*^. It is used for blood collection from newborns for metabolic studies and presents the benefits of having high conservation properties, requires small sample volumes, while remaining affordable and innocuous. The objective of the present study is to evaluate the analytical sensitivity of filter paper for *T*.*cruzi* qPCR detection.

## Material and Methods

### Sample preparation

A culture of VERO cells was used for this study. Upon reaching confluence, VERO cells were incubated with a culture of trypomastigotes forms of *T*.*cruzi* VD strain (Tc VI). (Brossas et al. 2017). The trypomastigotes were then allowed to invade VERO cells and after 72 hours, the parasites were recovered and counted in a Neubauer chamber. A suspension of 5 × 10^6^ tripomastigotes / mL was prepared

The suspension was diluted in blood (preserved in EDTA anticlotting agent) from non-infected donor to obtain concentrations between 50000 to 5 (50000, 5000, 4000, 3000, 2000, 1000, 500, 400, 300, 200, 100, 50, 5) parasites per milliliter of blood. Were aspirated aprox 75 µl of whole blood using a pipette with a disposable tip and was transferred the blood to the center of one circle without touching the filter paper directly with the tip of the pipette. The sample was loaded trying to fully saturate the circle. This procedure was repeat this procedure to fill all required circles of the card (Figure1.) An extra card was loaded with non-infected blood to be used as a negative control and an aliquot of the parasite dilution of liquid blood (5par/mL) was kept being used as a control of the extraction reaction. To dry the blood spots, were put the filter cards on a clean paper towel in a biohazard safety cabinet and let them dry. When the blood spots had a uniformly dark brownish color and no red areas are visible anymore, the drying process is complete. The cards were stored at room temperature and were processed between three and seven days after preparation.

**Figure 1.**
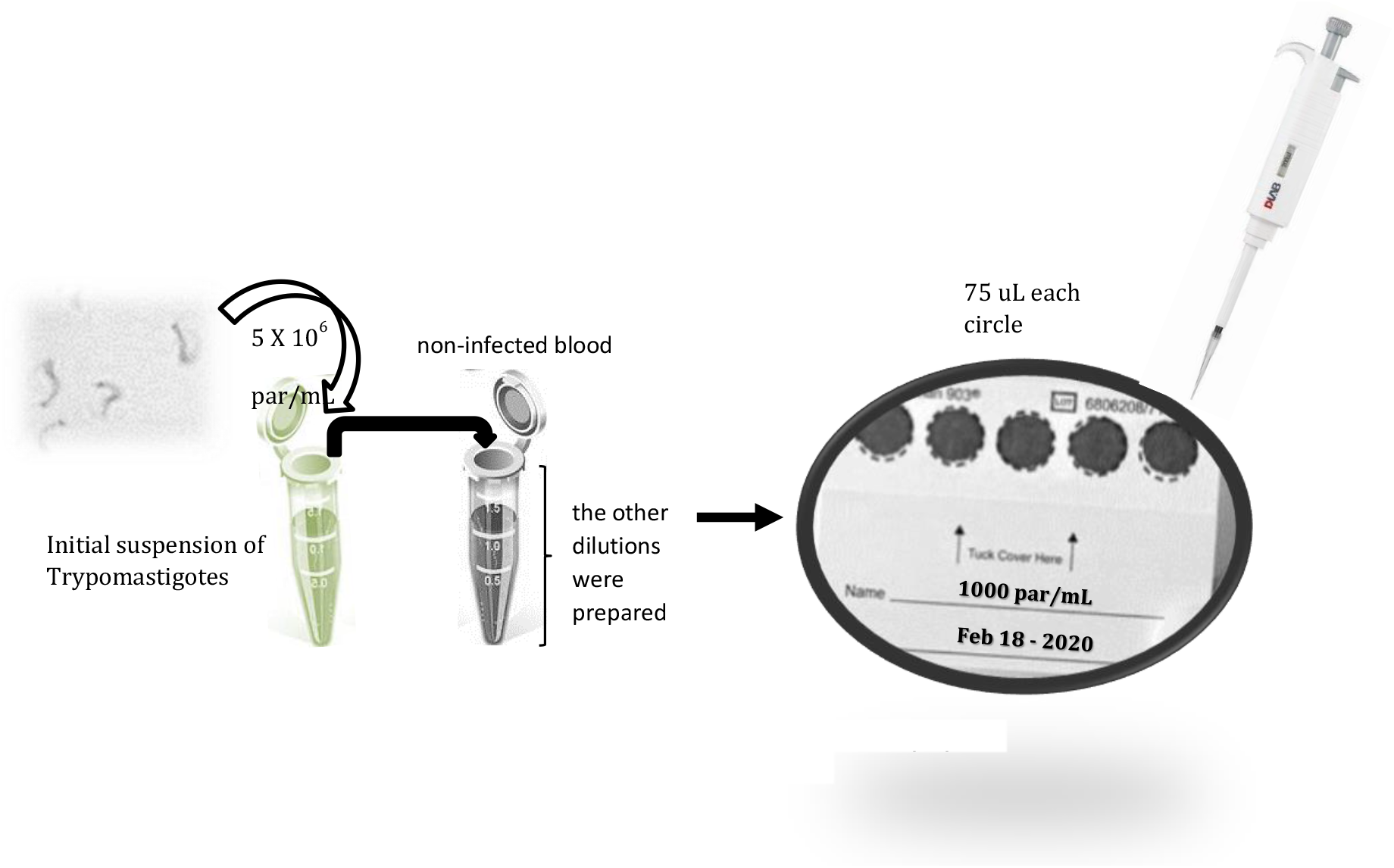
Sample preparation on Whatman 903 filter paper.

### DNA extraction

The DBS samples were lysed with ATL (sodium dodecyl sulphate) and DNAs were extracted using a Qiagen QIAamp DNA Mini Kit (Scanga et al., 2016 & Duffy el al. 2013). An internal amplification control IAC (ZErO-2 recombinant plasmid) plasmid gently provided by Dr Schijman and coworkers (INGEBI-CONICET Argentina) was included for the *T*.*cruzi*-PCR (Duffy et al 2009, Duffy et al. 2013). IAC amplification permitted to rule out false negative PCR results due to inhibitory substances or loss of DNA during sample processing. DNA was extracted from a whole spot (diameter, approximately 1 cm). The blood filter spot was transferred into a 2-ml tube. A single circle was processed for parasite concentrations above 1000 par/mL and two to five circles were processed for concentrations equal or less than 500 parasites/mL. The extractions were replicated to corroborate consistent results. The final DNA extract was stored at −18°C.

Two negative controls were included in the extraction: (1) blood for non *T. cruzi*-infected donors in Whatman 903 filter paper and (2) sterile water. Positive extraction control was made using anticoagulated blood infected with 5 *T*.*cruzi* parasites per milliliter (Figure 1) (Duffy et al. 2013).

### Quantitative PCR Assays

PCR was performed in 20 μL reaction mixtures containing DNA extracted from dried blood spot, 1X PCR FastStart Universal Probe Master Master Mix (Roche), 0.75mM SatDNA specific primers: cruzi1 (ASTCGGCTGATCGTTTTCGA), cruzi2 (AATTCCTCCAAGCAGCGGATA), 0.25 mM SatDNA specific probe cruzi3 (sonda) (CACACACTGGACACCAA) (Duffy et al. 2013) TaqMan™ MGB and water to ajust the final volume. Positive and negative controls were both processed in parallel with the different dilutions and were included in each run (Duffy et al. 2013). The specific primers for heterologous extrinsic control were:

IPCTqFw:5′-ACCGTCATGGAACAGCACGTA-3,

IPCTqRv:5′CTCCCGCAACAAACCCTATAAAT-3′IPC(sonda):5′VIC

AGCATCTGTTCTTGAAGGT 3′MGB-NFQ. TaqMan™ MGB Probe.

The qPCR products were analyzed using a LightCycler480 (Roche). The positive detection of parasitic DNA is based on the Cycle Threshold (Ct) value obtained for the qPCR.

## Results

*T*.*cruzi* DNA was detected on extractions at different dilutions from DBS: The limit of detection using just one DBS circle was 1000 parasites/mL (Table 1) and 400 parasites/mL using two DBS circles (Table 2.)

**Table I.**
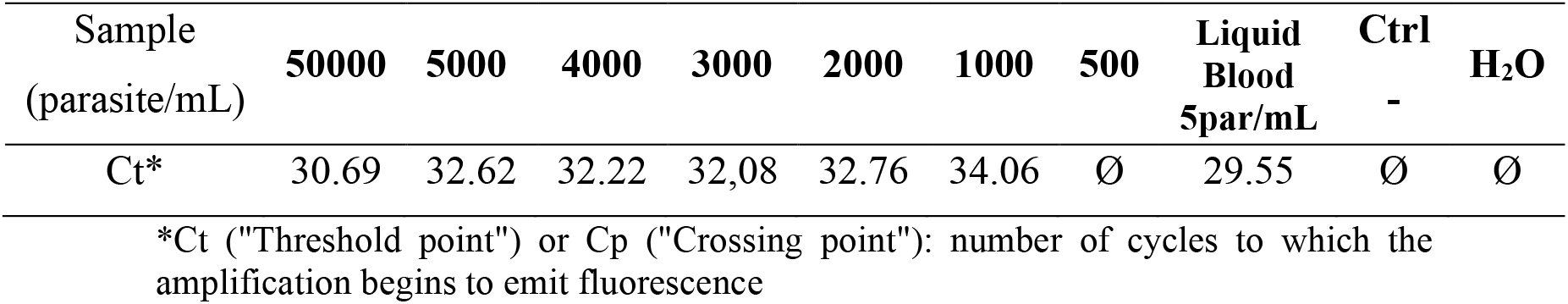
*T. cruzi* DNA detection through qPCR for DBS extraction using **one** DBS circle

**Table II.** *T*. cruzi DNA detection through qPCR using two DBS circle.

| Sample<br>(parasite/mL) | 500 | 400 | 300 | 200 | 100 | 50 | Liquid<br>Blood<br>5par/mL | Ctrl<br>+ | H<sub>2</sub>O |
| --- | --- | --- | --- | --- | --- | --- | --- | --- | --- |
| Ct | 32.75 | 33.95 | 35< | 35< | Ø | Ø | 29.75 | Ø | Ø |

As indicated in table II, using two DBS circles, a signal was observed among those with 500 parasites/mL to 400 parasites/mL with Ct 32.75 and 33.95 respectively, however the concentrations of 300 and 200 parasites/mL also gave a signal although it was very late with a Ct = 35 (Table II).

Extractions with four and five DBS circles were tested for concentrations of 5, 50, 100, 200 and 300 parasites per milliliter but no fluorescence signal was visible.

Figure 2 shows the inverse relationship between Ct and DNA concentration in ng/µL.

**Figure 2.**
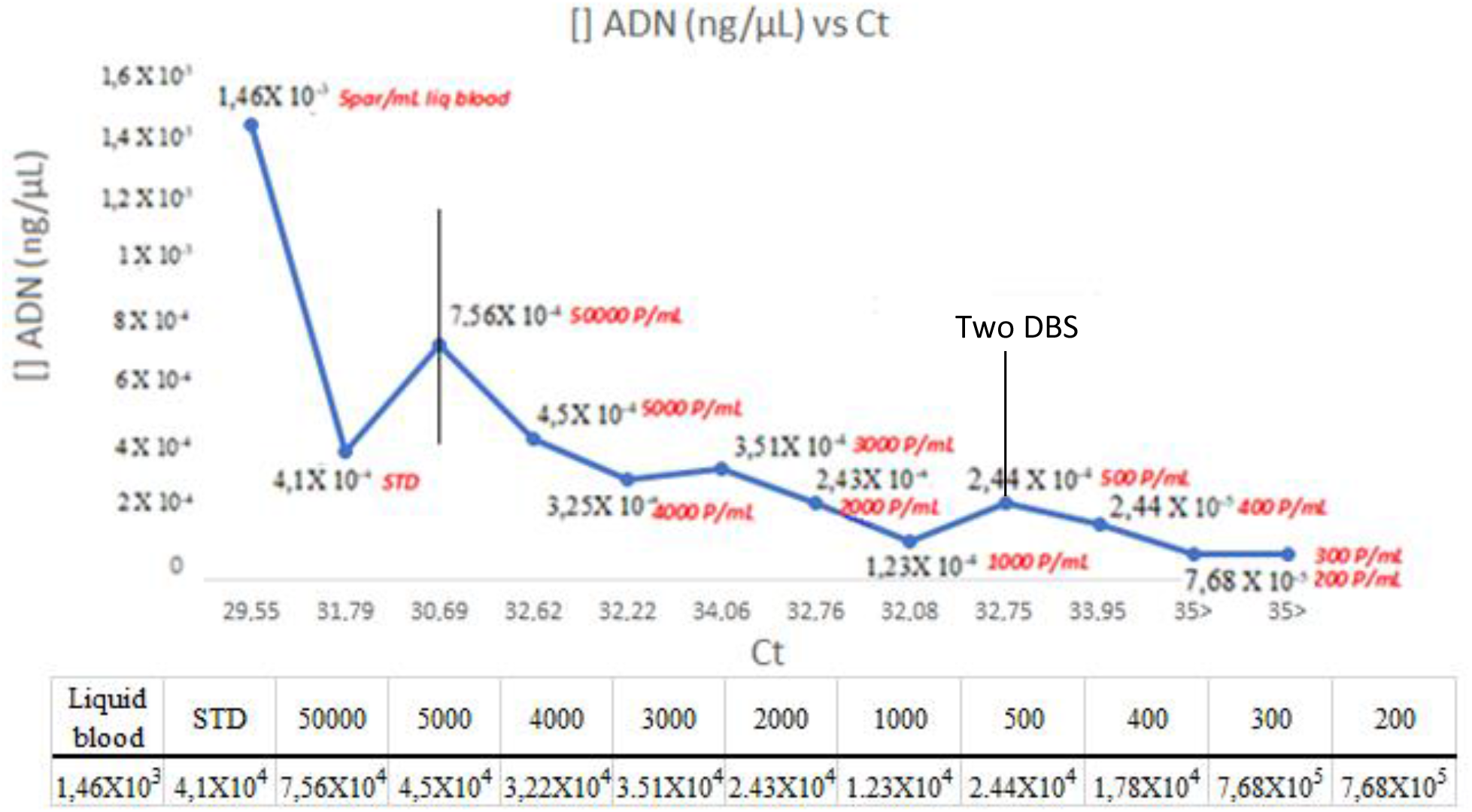
relationship between Ct and DNA concentration in ng/µL and parasite dilution/mL.

## Discussion

The conservation of dried blood samples on filter paper is a proven successful method for the PCR detection of DNA from other parasitic infectious disease agents, such as Plasmodium for malaria diagnosis. Several assays have also been made for the detection of congenital cytomegalovirus (CMV) infection and HIV infection (Viana et al. 2010, Lorenzana de Rivera & Murillo 2003, Oriane et al. 2008, Sacanga et al. 2006).

In this study, it was possible to obtain *T. cruzi* DNA from samples preserved in filter paper, proving that the parasite’s DNA was effectively protected from degradation (Table I). It requires a small amount of blood, facilitating sample collection, particularily with newborns, and allows storage during long periods of time, without toxic preservative reagents (Maeno et al. 2008, Viana et al. 2010, Vitale et al. 2018).

Direct parasitological detection using the microhematocrit test (MH) is the diagnostic methods of choice for newborns (Carlier et al. 2019). MH is based on the observation of parasites (trypomastigotes) in the white blood cell layer after centrifuging blood in a heparinized capillary tube. The MH technique is recommended for diagnosis of CD in infants because it uses small volumes and detects between 40 and 1000 parasites /mL (Freilij el at. 1983, Torrico et al. 2005), but it requires skilled and highly trained operator which may lead to variability in diagnostic sensitivity across centers. MH needs an anticoagulated fresh sample to enable detection of motile trypomastigote forms (Carlier et al. 2019, Bern et al. 2011, Altcheh & Freilij 2019).

Several published studies show that PCR detection of *T. cruzi* gives a better detection limit (0.5-1 parasites / milliliter) (Duffy et al. 2013) compared to the MH technique (40- 1000 parasites / milliliter) (Freilij el at. 1983, Torrico et al. 2005). However, in all cases of PCR detection, this methodology requires that the sample is preserved in a toxic reagent such as guanidine (Schijman et al. 2011, Duffy et al. 2013, Ramirez et al. 2015, Coronado 2016, Rivero et al. 2017)

The proposed method has a minimum detection of 400 parasites/mL of blood, on filter paper (Table II); although this is an intermediate sensitivity, this technique can be widely applied in primary care centers far from high-tech centers and counts several advantages: the properties of the samples are conserved, dry blood on the filter paper is not infectious and biologically stable, allowing the long term storage of the samples without losing or deteriorating DNA, and do not require refrigerated conditions or toxic preservatives agents anymore (Viana et al. 2010, Vitale et al. 2018).

Considering the utility of Whatman filter paper in the analysis of other congenital diseases and in response to the need of establishing early diagnosis on congenital CD in Argentina, this method constitutes an opportunity for the detection of *T. cruzi* DNA by facilitating the transport and the handling of the samples (Vitale et al. 2018). The implementation of this technique in Argentina implies a technological development that would allow the diagnosis of newborns and will have a great impact, especially in populations far from large cities, allowing the collection and subsequent transport of samples to laboratories for processing.

Additional testings on the feasibility of using filter paper for the conservation and transport of samples, beforemolecular analysis purposes have been conducted (Perozo et al. 2006, Aysanoa et al. 2017, Vitale et al. 2018, Rivero et al. 2018). However FTA filter paper was used, which is more expensive and not widely used, in the opposite of Whatman 903 paper which is standardized in national public health programs for the detection of metabolic diseases (msal.gob.ar 2011)

This study attempts to provide an approach to a useful and necessary alternative for the collection and conservation of samples for congenital CD studies. However, in orderto ensure a complete diagnosis by combining parasitological, immunological and molecular techniques, further studies should be conducted to ensure proper validation and standardization. Clinical trials of samples preserved on Whatman 903 filter paper should be conducted to determine their potential usefulness and performance in the diagnosis of CD in the context of neonatal research and transfer this technology to be used for diagnosis of Congenital CD in the future.

## Data Availability

All relevant data are within the manuscript and its Supporting Information files.

## Footnotes

1 (http://www.msal.gob.ar/images/stories/bes/graficos/0000000068cnt-p01-manual-de-procedimiento.pdf).

## Notes

### Competing Interest Statement

The authors have declared no competing interest.

### Funding Statement

This work was funded by financial support of the Hospital de Ninos Ricardo Gutierrez

### Author Declarations

The Ethics Committee and Review Board of the Hospital de Ninos Ricardo Gutierrez approved this study

